# Subtle language deficits in WAB-recovered patients at 12 months after left-hemisphere stroke

**DOI:** 10.64898/2026.06.19.26356022

**Authors:** Manuel Jose Marte, Mathew Chaves, Lindsey Kelly, Isidora Diaz-Carr, Voss Neal, Andreia V. Faria, Melissa D. Stockbridge, Argye E. Hillis

## Abstract

**Background:** The Western Aphasia Battery-Revised (WAB-R) Aphasia Quotient is the most widely used standardized post-stroke aphasia measure; its conventional 93.8 cutoff has limited sensitivity to mild residual impairment. Beyond the cutoff, it offers limited objective discourse assessment, no action-naming assessment, and naming tests limited to very common objects.

**Aims:** We sought short tests capturing subtle aphasia in patients recovered above the WAB-R threshold, then its demographic and lesion correlates.

**Methods & Procedures:** Sixty-seven patients with acute left-hemisphere ischemic stroke completed acute structural MRI and a 12-month language battery comprising the WAB-R, Boston Naming Test (BNT), Hopkins Action Naming Assessment (HANA), and Modern Cookie Theft (MCT) picture description. Hierarchical logistic (binary deficit) and linear (control-referenced composite z-score) regressions evaluated acute aphasia history, sex, education, age, acute depression (PHQ-9), and residualized regional lesion load.

**Outcomes & Results:** Of 67 participants, 45 (67%) recovered above the WAB-R threshold. Of these, 18 (40%) had residual deficits on at least one supplemental test (“subtle aphasia”). BNT plus MCT content-unit count captured all 18 (100%); HANA added none beyond these two. The binary model discriminated deficit from no-deficit at AUC = 0.80 (95% CI [0.70, 1.00]); higher education significantly lowered deficit odds (OR = 0.80/year, 95% CI [0.64, 1.00], p = .049). On the continuous composite, acute PHQ-9 independently predicted 12-month outcome (β = −0.13 per point, 95% CI [−0.22, −0.04], p = .006, cumulative R² = 0.38). Applying the Senthilkumar et al. (2026) stricter cutoff (WAB-AQ ≥ 96.7) reclassified 12 of 45 (27%) out of recovery, capturing 8 of 18 (44%) subtle-aphasia patients. Composite residualized lesion load did not differentiate the groups when adjusted.

**Conclusions:** Above the WAB-R recovery threshold, subtle aphasia is present on the BNT or MCT in ∼40%, with higher education associated with lower odds at 12 months; acute depression emerged as a candidate correlate but did not survive removal of a single high-influence observation, warranting replication in larger samples. Regional lesion variables informative at greater stroke severity contribute little as large lesions cluster in the persistently aphasic group, reducing lesion variance within the recovered subgroup and its discrimination of subtle deficits. This adds to evidence that clinicians should not infer complete language recovery from the WAB-AQ alone, and that identifying residual deficits may require greater investment in behavioral assessment and consideration of alternative WAB-AQ cutoffs. Structural anatomical information, by contrast, appears to add little discriminative value at the upper performance range.

## 1. Introduction

The Western Aphasia Battery-Revised (WAB-R; Kertesz, 2012) is one of the most widely used standardized assessments for post-stroke aphasia, and the WAB-R Aphasia Quotient (AQ) ≥ 93.8 cutoff classifies a patient as “non-aphasic.” A recent systematic review documented over four decades of WAB-R use across research and clinical settings, with the AQ continuing to be the most frequently reported summary measure for characterizing aphasia severity and recovery in the field (Kertesz, 2022). Yet a substantial body of work has documented that patients above this threshold can present with persistent communicative difficulty (Cavanaugh & Haley, 2020; Cunningham & Haley, 2020; Dalton & Richardson, 2015; Fromm et al., 2017; Kurland et al., 2026), creating a classification gap with clinical stakes, as patients above cutoff may be denied access to rehabilitation services despite measurable residual impairment (Kurland et al., 2026). This gap motivates an empirical question: how prevalent are residual deficits above the WAB-AQ threshold, and what patient-level variables may predict them?

Aphasia at the threshold of clinical detection has been recognized for over half a century. In medical usage, the term “latent” typically refers to a condition that is present but not currently active or manifesting (as in latent tuberculosis or latent virus); the deficits described here, by contrast, are actively manifesting on behavioral measures and simply fall below the detection threshold of the WAB-AQ at conventional cutoffs. We therefore use “subtle aphasia” throughout, including in our description of the historical and contemporary literature that has used the term “latent aphasia” for this construct (Boller, 1968; Pichot, 1955). Indeed, Pichot (1955) introduced the concept of subtle aphasia in the context of arteriosclerotic dementia, though Boller (1968) extended the construct to non-aphasic left-hemisphere stroke survivors, demonstrating that subtle linguistic impairment which is not detected upon superficial examination can be evinced by additional testing. Boller (1968) compared 22 left-hemisphere and 37 right-hemisphere brain-damaged patients without clinical aphasia against 20 controls, finding that the subtle aphasia effect was demonstrated on the Token Test, where left-hemisphere patients performed below right-hemisphere patients on auditory comprehension. Expressive measures did not separate the hemispheric groups in Boller’s data, and Boller called for more sensitive tests as the methodological gap for future work.

Contemporary work has revived and advanced the construct (e.g., DeDe & Salis, 2020; Fromm et al., 2017, 2024; Gilman & LaCroix, 2026; Laks et al., 2025, 2026; Salis et al., 2021; Stark et al., 2024) and converges on a single empirical pattern across various behavioral measures, which is that stroke survivors who score above the diagnostic cutoff on standardized aphasia batteries differ measurably from neurotypical controls on tasks of lexical retrieval and processing, connected discourse, and syntactic comprehension.

Discourse-level evidence forms the largest body of contemporary work. For example, Fromm et al. (2017), in 28 chronic non-aphasic-by-WAB (NABW) patients, demonstrated reduced words per minute, lexical diversity, and main concept production relative to controls, with the authors suggesting that connected speech, in imposing minimal external scaffolding for lexical access and discourse organization, helps to surface residual deficits in these domains relative to structured assessments. Similarly, Fromm et al. (2024) analyzed an AphasiaBank corpus of 281 persons with aphasia and 257 controls, of whom 31 (11%) were classified as NABW per Fromm et al.’s (2017) criteria, and found that PWA (including the NABW group) produced significantly fewer total words, fewer words per minute, and more pauses, repetitions, revisions, and phonological fragments than controls. Next, Stark et al. (2024) replicated reduced core lexical access and slower retrieval across three discourse tasks in 29 TalkBank subtle aphasia speakers relative to 30 mild cognitive impairment speakers and 56 cognitively healthy adults, pointing to a time-sensitive component of lexical-semantic access that is impaired in subtle aphasia. Further, Cunningham and Haley (2020) examined connected speech samples from 478 speakers in AphasiaBank, including 28 stroke survivors classified as NABW, 225 individuals with aphasia, and 225 neurotypical controls; they compared two automated indices of lexical diversity, the moving-average type-token ratio (MATTR-5), which computes type-token ratios within a sliding fixed-length window across the sample, which distinguished the NABW group from controls. The authors attributed this sensitivity to the local nature of the MATTR’s detection, that is, by analyzing word repetition within small windows, the MATTR-5 captures local lexical repetition that arises from failures of lexical retrieval, speech production difficulties, or repetitive morphosyntactic structures stemming from subtle grammatical deficits - repetition that a length-invariant global lexical-diversity measure does not consistently elicit. Together, these studies establish that the production of connected speech in the WAB-recovered range carries a measurable group-level deficit.

Next, individual classification evidence uses brief sensitive measures to assign deficit status to single patients, and these studies converge on the conclusion that brief naming and sentence-level instruments can identify residual deficits in WAB-recovered patients at the single-case level. Laks et al. (2025), in 27 NABW patients matched to 68 controls, found that combining FAS letter fluency with two-letter fluency (Po, Ta) achieved 89% sensitivity and 81% specificity for individual-case detection in approximately 11 minutes, though category fluency did not reach the .70/.70 threshold. The authors attributed this sensitivity to the speed component of lexical retrieval, in that rapid lexical access stresses lexical-semantic systems that may remain mildly compromised after stroke, even when patients can produce the same items given more time. Recall Fromm et al. (2024) and Stark et al. (2025)’s findings with respect to timing-and-processing-based differences: Laks et al. (2025) provided direct evidence that the first 30 seconds of FAS letter fluency carried the highest individual-test AUC (.90) of any measure in their battery, higher than total trial output (.87) or the second 30 seconds (.79). The authors suggested that it was consistent with the view that the earliest, most-accessible-items phase of retrieval stresses lexical-semantic access most steeply. Laks et al. (2026) then disentangled the underlying cognitive components, showing that deficits in rapid lexical retrieval (timed picture naming and lexical-retrieval items on Antelopes and Cantaloupes), rather than non-lexical executive control, drove the NABW-control letter-fluency gap. Next, Walker et al., (2022) developed the Severity-Calibrated Aphasia Naming Test (SCANT), a 20-item naming instrument developed a large mixed-severity calibration sample to span a wider difficulty range than conventional confrontation naming tests. In a validation sample of 83 PWA and 20 controls, the SCANT correctly classified 92.2% of individuals and outperformed the full set of naming items for predicting aphasia severity, supporting the brief-instrument-with-mild-range strategy that connects this evidence stream to the discourse strand, i.e., both lines of work establish that what fails at the WAB-AQ ceiling is sensitivity to mild residual impairment, which different instruments recover through different mechanisms (item-difficulty calibration in naming; rapid-retrieval timing in fluency; sustained discourse demands in connected speech). Lastly, Gilman and LaCroix (2026) showed, in 64 left-hemisphere stroke survivors, that performance on a multi-sentence comprehension paradigm differentiated subtle aphasia from controls and from anomic aphasia in terms of accuracy on syntactically complex sentences (specifically, object-relative constructions), with the authors suggesting that the syntactic-complexity gradient implicates residual sentence-processing deficits that single-word and simple-sentence tasks miss because they do not sufficiently tax the cognitive resources required for integration of non-canonical syntax.

Beyond the discourse-level and brief-instrument evidence reviewed above, complementary methodological streams have established subtle-aphasia detection through subjective report and through measures of cognitive processes. Cavanaugh and Haley (2020) conducted semi-structured qualitative interviews with five individuals classified as having very mild aphasia (WAB-AQ above the recovery cutoff), and all five described being challenged across multiple aspects of communicative life participation, with consistent reports of needing more preparation for communication, slower response times, residual anomia, and increased communicative effort relative to pre-stroke. The authors interpreted these qualitative accounts as evidence that the WAB-R underestimates the functional communicative experience of recovered patients, supporting the position that subjective accounts of difficulty are a valuable complement to the WAB-AQ when classifying recovery. Silkes et al. (2021) extended the evidence in a different direction, administering the Temple Assessment of Language and Short-term Memory in Aphasia (TALSA) to 7 individuals with subtle aphasia and 24 neurotypical controls; 21 of 40 TALSA subtests differentiated the subtle-aphasia group from controls, with all of these subtests engaging verbal short-term memory and some additionally engaging working memory, establishing that residual impairment in this range is detectable not only at the linguistic surface but also in the verbal short-term and working memory substrates that support language processing.

However, detection depends critically on the measurement instrument and the cutoff it imposes. These discourse- and individual-classification findings raise the question as to whether the WAB-R cutoff value itself and the way it is operationalized are adequate for capturing the residual deficits these measures elicit. Senthilkumar et al., (2026) directly tested the diagnostic sensitivity of the conventional 93.8 cutoff in 171 acute ischemic stroke patients within 2 weeks of symptom onset, comparing WAB-R classification against discourse-based aphasia identification on Cookie Theft and Modern Cookie Theft picture descriptions; 33 of 171 patients (19%) showed discourse-level aphasia despite a non-aphasic WAB-R score, and the authors proposed a revised cutoff of WAB-AQ ≥ 96.7 to improve detection through their receiver operating characteristic analysis. Similarly, the aforementioned work of Walker et al. (2022) and Laks et al. (2025; 2026) provided complementary tools showing that brief sensitive measures retain discriminative accuracy at the WAB-AQ ceiling where conventional naming tests reach floor on errors.

The instrument and cutoff questions reviewed above frame what counts as residual impairment in the recovered range, but a separate question is which patient-level variables may locate it. Predictors of post-stroke aphasia outcome have been characterized extensively across the full severity spectrum (for a review, see (Marte et al., 2025); for an individual-participant data meta-analysis of n = 5,928 patients, see Ali et al., 2021). Initial severity is the strongest single predictor of chronic outcome (Ali et al., 2021; Lazar et al., 2010). Beyond severity, gross lesion volume contributes additional variance (Hillis et al., 2018; Wilson et al., 2023); damage to canonical left-hemisphere language regions predicts domain-specific impairment in naming, discourse, and comprehension (e.g., Baldo et al., 2013; Wilson et al., 2023). Relatedly, factors like educational attainment likely relate to pre-stroke cognitive resources that buffer the functional expression of brain injury (Ali et al., 2021; González-Fernández et al., 2011; Stern, 2009, 2012); and acute depression is bidirectionally linked to language outcome (Hackett & Pickles, 2014; Hilari et al., 2012; Robinson & Jorge, 2016).

In sum, the present study examined the prevalence and correlates of subtle aphasia at 12 months after left-hemisphere ischemic stroke. We used additional expressive language measures to identify residual deficits in WAB-recovered patients, and also evaluated the contribution of clinical history, demographic, mood, and structural anatomical variables to deficit status within the WAB-recovered subgroup and tested whether residualized regional lesion damage discriminates recovered patients with from those without residual deficits. The evidence reviewed above motivates three hypotheses. First, (H1) residual deficits (subtle aphasia) will be detectable in a substantial minority of WAB-recovered patients. Secondly, (H2) demographic and lesion volume (but not regional lesion load) will differentiate between those with subtle aphasia versus no deficit, among WAB-recovered patients. Lastly, (H3), the application of the Senthilkumar et al. (2026) alternative recovery cutoff of WAB-AQ ≥ 96.7 will capture some of the WAB-recovered patients with subtle aphasia, indicating that an alternative aphasia threshold can address the upper-end sensitivity problem.

## 2. Methods

### 2.1 Participants

Participants were drawn from an ongoing longitudinal program at Johns Hopkins Hospital that has enrolled 617 English-speaking adults with acute left-hemisphere (LH) ischemic stroke since 2007. Inclusion criteria for the present analysis were a confirmed LH ischemic stroke on magnetic resonance imaging (MRI); acute WAB-R assessment within the first week post-onset (median 2 days, mean 2.8 days, SD = 2.2); completion of 12-month follow-up; and availability of a complete neuroimaging processing pipeline. Patients were excluded from longitudinal monitoring at enrollment if they had stroke limited to the brainstem or cerebellum, hemorrhagic stroke, preexisting neurological disease affecting language or cognition (e.g., dementia), intellectual disability, or severe or uncorrected vision or hearing loss. Because the core language battery and discourse protocol were updated across the enrollment window (the WAB-R replaced earlier diagnostic protocols as the standard aphasia assessment partway through the program, and the picture-description task used to derive discourse measures was added at a later phase) paired acute and 12-month WAB-AQ were available for an analytic subset of the full enrollment, and discourse measures for a smaller subset. All analyses in the present study used the harmonized measures available across enrollment phases. The analytic sample for the present study comprised 67 patients with all required language and imaging measures. Of these, 30 were never aphasic by the WAB-R at either acute or 12-month assessment, 15 had aphasia acutely that resolved by 12 months, and 22 had persistent aphasia at 12 months; the 30 never-aphasic and 15 resolved patients together formed the WAB-recovered analytic subgroup (n = 45). Within-subgroup demographic and clinical characteristics for the WAB-recovered analytic cohort, separated by residual-deficit status, are reported in Table 1; sample characteristics for the full N = 67 cohort by 12-month WAB-AQ recovery group (never aphasic / resolved aphasia / persistent aphasia) are reported in Supplementary Table S1. All study procedures were approved by the Johns Hopkins University School of Medicine Institutional Review Board (protocol NA_00042097); written informed consent was obtained from participants or their legally authorized representatives, in accordance with the Declaration of Helsinki, prior to participation.

**Table 1.** Sample characteristics within the WAB-recovered subgroup, separated by residual subtle-aphasia status.

| Variable | No deficit ($n = 27$) | Subtle aphasia ($n = 18$) | Test |
| --- | --- | --- | --- |
| Age, years | 59.03 (12.55) | 61.06 (13.37) | $t(34.9) = 0.51$, $p = .614$ |

| Variable | No deficit (n = 27) | Subtle aphasia (n = 18) | Test |
| --- | --- | --- | --- |
| Female sex, n (%) | 11 (41) | 5 (28) | $\chi^2(1) = 0.33, p = .567$ |
| Education, years | 15.52 (2.74) | 13.78 (3.28) | $t(31.9) = -1.86, p = .072$ |
| Days post-onset | 2.8 (1.9) | 3.0 (2.2) | $t(29.1) = 0.31, p = .762$ |
| Acute WAB-AQ | 91.60 (13.98) | 86.39 (23.68) | $t(24.9) = -0.84, p = .408$ |
| Log lesion volume | 8.17 (1.82) | 8.19 (1.59) | $t(39.0) = 0.04, p = .970$ |
| Acute PHQ-9 | 3.32 (4.01) | 5.47 (4.16) | $t(28.8) = 1.60, p = .120$ |
| SLP sessions 0-12 mo. | 15.96 (24.92) | 12.77 (15.69) | $t(34.0) = -0.48, p = .637$ |
| Composite z | 0.44 (0.62) | -1.00 (0.85) | $t(28.8) = -6.17, p < .001$ |

*Note. M (SD) or n (%). Welch two-sample t-tests for continuous variables; Pearson chi-square for sex. Composite z is the equally weighted mean of standardized BNT, HANA, and MCT content units; deficit and no-deficit groups separate by construction.*

### 2.2 Language assessments and recovery classification

The 12-month language battery included the WAB-R, the Boston Naming Test (BNT; Kaplan et al., 2001), the Hopkins Action Naming Assessment (HANA; Breining et al., 2022), and the Modern Cookie Theft (MCT) picture description (Berube et al., 2019). The WAB-R was administered acutely as well as at 12 months. WAB-recovered status at 12 months was defined as WAB-R AQ ≥ 93.8 per Kertesz (2012). Within the recovered subgroup, residual deficits were defined as below-cutoff performance on at least one of three additional language measures: the BNT, the HANA, or the MCT content-unit count. Missing measure values were treated as not flagged. BNT cutoffs were the education-corrected normative cutoffs of Jefferson et al., (2007). The HANA cutoff was the impairment cutoff applied by Breining et al. (2022), corresponding to a raw score of 18 or lower. The MCT content-unit cutoff was 22, corresponding to one standard deviation below the healthy control mean reported by Berube et al. (2019).

We separately examined speech-language therapy dose between acute and 12-month assessments, which was operationalized as the number of speech-language therapy sessions documented in the medical record during the 0-12-month post-stroke window. Age at stroke and years of education were recorded at enrollment. Acute aphasia history was operationalized as a binary indicator of whether the patient had been classified as aphasic by the WAB-R at the acute assessment (resolved = 1, never aphasic = 0). The acute PHQ-9 (Patient Health Questionnaire-9; Kroenke et al., 2001) was administered as part of the acute assessment battery to index acute self-reported depressive symptoms.

### 2.3 Neuroimaging

Baseline imaging was acquired at the time of admission for the index stroke on a 3 Tesla scanner; all patients underwent diffusion-weighted imaging, from which ischemic lesion volume was derived. Acute diffusion-weighted MRI sequences were processed with the Acute-stroke Detection Segmentation (ADS) pipeline (Faria, 2021), which generated an initial lesion mask via deep-learning segmentation on diffusion-weighted imaging. Masks were reviewed and manually corrected in ITK-SNAP (Yushkevich et al., 2016) by a trained research assistant (M.C.) blinded to 12-month outcome and subsequently verified by the lead author (M.J.M.). Proportional damage to each cortical parcel of the Brain Parcellation Map (BPM; (Faria et al., 2012; Oishi et al., 2009) was computed via ADS.

Ten left-hemisphere cortical regions were selected a priori from the BPM atlas based on convergent evidence from lesion-symptom mapping (Thye & Mirman, 2018) and the dual-stream model of language (Hickok & Poeppel, 2007). Eight regions fall within canonical left perisylvian language territory with documented lesion-behavior associations to production, comprehension, or repetition: the inferior frontal gyrus pars opercularis (BA 44) and pars triangularis (BA 45) for speech production and syntactic processing (Friederici & Gierhan, 2013); the superior temporal gyrus for auditory comprehension via the ventral phonological-recognition pathway (Mirman & Thye, 2018); the supramarginal gyrus and angular gyrus for phonological and lexical-semantic processing (Mirman & Thye, 2018); the middle temporal gyrus and inferior temporal gyrus for lexical-semantic retrieval (Mirman & Thye, 2018); and the insula for speech motor planning (Mirman & Thye, 2018). Two extra-perisylvian frontal regions were included to test contributions outside the core perisylvian network: the middle frontal gyrus, given its role in executive-control aspects of connected-speech production (Mirman & Thye, 2018), and the superior frontal gyrus, as the cortical origin of the frontal aslant tract supporting speech initiation and verbal fluency, with associations to complex syntactic processing. Subcortical structures (thalamus, putamen) were excluded from the cortical-parcel feature class to avoid conflating distinct tissue types and due to absence of significant subcortical involvement in all patients of interest. Each region’s lesion load was residualized on log total lesion volume via linear regression fit within the WAB-recovered analytic subgroup (DeMarco & Turkeltaub, 2018), isolating the relative pattern of regional damage from total lesion extent. A composite was computed as the sum of residualized loads across the ten parcels.

### 2.4 Continuous composite z-score

The continuous outcome was a control-referenced composite z-score, constructed as the equally weighted mean of standardized scores on the three measures:

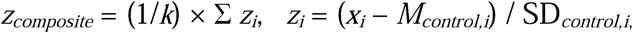

where *k* is the number of available measures for the patient, *x□* is the patient’s raw score on measure *i*, and *M_control,i_*and SD*_control,i_* are the control mean and standard deviation for measure *i*. As an illustrative example, a patient with HANA = 17, BNT = 21, and MCT content units = 20, given published control means (SDs) of 23.9 (3.46) for HANA (Breining et al., 2022), 26.3 (2.48) for BNT (Breining et al., 2022), and 34.7 (11.6) for MCT content units (Berube et al., 2019), would have a composite z-score of approximately −1.80 (*z_composite_* = [(17 − 23.9)/3.46 + (21 − 26.3)/2.48 + (20 − 34.7)/11.6] / 3 = [(−1.99) + (−2.14) + (−1.27)] / 3 = −5.40 / 3 ≈ −1.80). The composite z was computed as the mean of available z-scores for each patient, with k ranging from 1 to 3, yielding a base sample of 45 recovered patients. A higher composite z indicates better performance.

### 2.5 Statistical analyses

All analyses were conducted in R version 4.3.0 (*R: The R Project for Statistical Computing*, n.d.). Continuous variables in descriptive comparisons were tested with two-sample t-tests using Welch’s correction for unequal variances; categorical variables were tested with Pearson chi-square, and confidence intervals on proportions were computed using the Wilson interval; all other 95% confidence intervals were Wald unless otherwise noted.

We examined the differential contribution of each of the three additional language measures to overall subtle deficit identification. For each WAB-recovered patient with a deficit on at least one of the supplemental tests (n = 18), we coded which of the three measures (BNT below the education-corrected cutoff, HANA ≤ 18, MCT content units < 22) were positive, treating missing measure values as no deficit. We then enumerated the seven non-empty subsets of the three measures and, for each subset, computed sensitivity for detecting subtle deficits as the proportion of these 18 patients positive on at least one measure in the subset.

Two parallel hierarchical regression analyses evaluated correlates of residual deficit status. The binary outcome was the deficit indicator (deficit on any of the three additional language measures). The continuous outcome was the control-referenced composite z-score described in 2.4.

The continuous composite z-score complements the binary indicator: where the binary outcome reflects a clinical-classification decision (deficit vs. no deficit), the continuous z-score uses the full graded variation in performance across the three measures, providing the inferential power that the binary indicator loses to dichotomization (MacCallum et al., 2002).

Both regression models followed the same hierarchical step ordering: Step 1 included acute aphasia history (binary, resolved = 1) and sex (binary, female = 1); Step 2 added years of education; Step 3 added age at stroke; Step 4 added composite residualized lesion load; Step 5 added acute PHQ-9 score. PHQ-9 was placed at Step 5 as the most distal step because it is a behavioral marker assessed within days post-onset that is plausibly downstream of both demographic factors and the structural lesion; the comparison between Step 4 and Step 5 isolates the depression contribution above and beyond the structural model.

For the binary outcome, hierarchical logistic regression yielded odds ratios with 95% confidence intervals at each step, likelihood-ratio tests against the previous nested step, and discriminative accuracy summarized by area under the receiver operating characteristic curve (AUC). AUC values range from 0.5 (no discrimination) to 1.0 (perfect discrimination); values of 0.70, 0.80, and 0.90 are conventionally interpreted as acceptable, good, and excellent discrimination (Hosmer & Lemeshow 2000). Confidence intervals on AUC were obtained by bootstrap percentile resampling with 2,000 replicates as implemented in the pROC package (Robin et al., 2011). When the analytic *n* decreased between steps because of covariate missingness, the previous step’s model was refit on the smaller subgroup before the likelihood-ratio comparison. For the continuous outcome, hierarchical linear regression yielded standardized betas with 95% confidence intervals, model R², and F-change tests at each step.

Acute language scores on each of the three measures were correlated with the 12-month continuous composite as convergent construct evidence; bivariate Pearson correlations are reported in 3.4.3. Speech-language therapy dose between acute and 12-month assessments was tested as a single bivariate correlate of both binary and continuous outcomes to address possible treatment confounding, also reported in 3.4.3.

For the regional lesion-discrimination question, composite residualized regional lesion load was compared between deficit and no-deficit recovered patients with a t-test on the mean difference.

## 3. Results

### 3.1 Prevalence

Among the 45 WAB-recovered patients, 40% (18 of 45, 95% CI [27%, 55%]) showed a residual deficit on at least one standardized test. Application of the Senthilkumar et al. (2026) alternative recovery cutoff of WAB-AQ ≥ 96.7 reclassified 12 of 45 WAB-recovered patients (27%) out of recovered. Of the smaller recovered-at-≥96.7 subgroup (n = 33), 10 patients (30%) had a residual deficit. Figure 1 shows the deficit-classification logic alongside prevalence.

**Figure 1.**
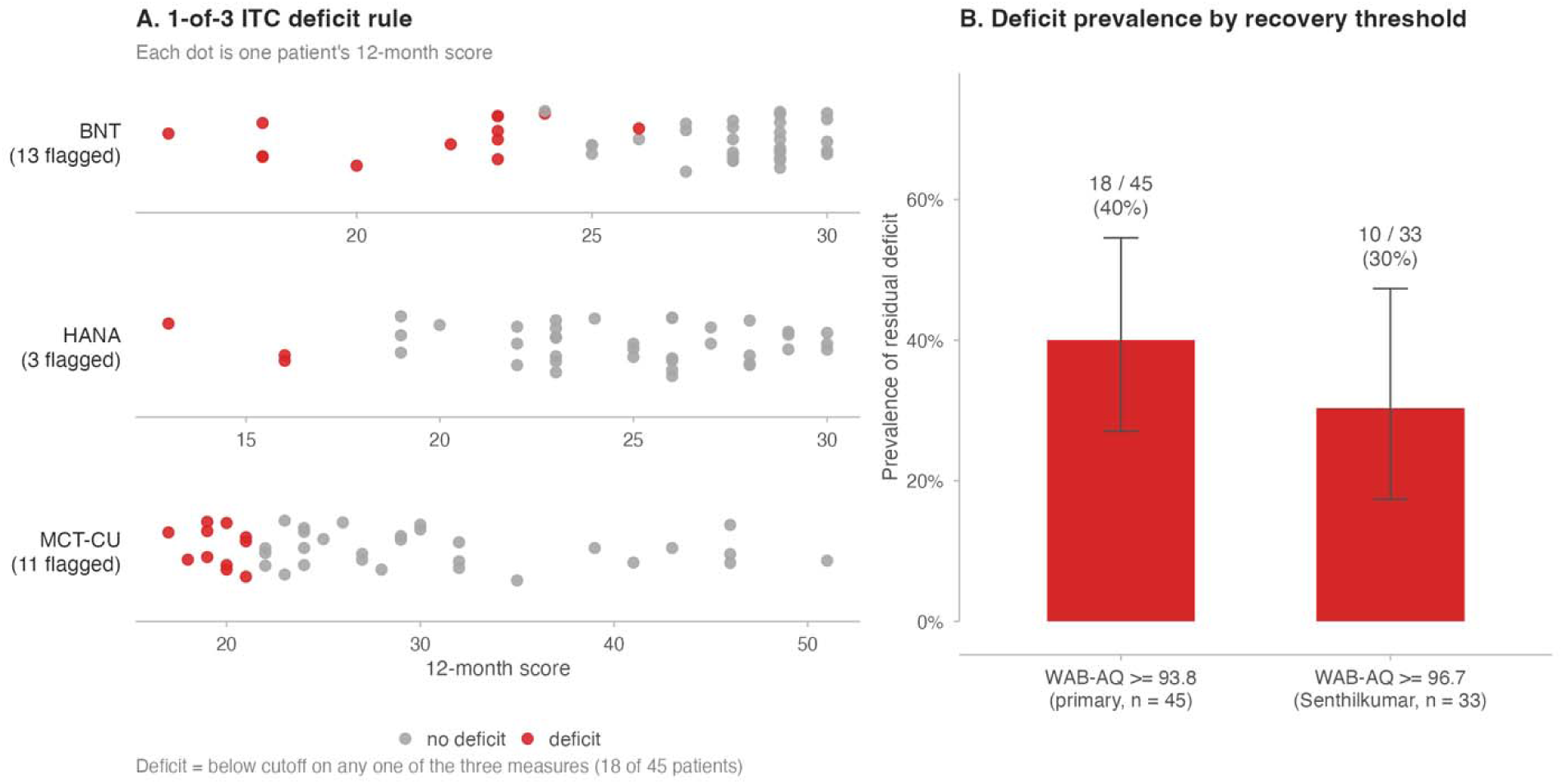
Residual language deficit classification and prevalence within the WAB-recovered subgroup. (A) 12-month score for every recovered patient on each of the three deficit measures (Boston Naming Test, HANA, and MCT content units); each dot is one patient. Red dots are below the measure-specific cutoff (a deficit on that measure); grey dots are at or above. A patient is classified as having a residual deficit if any one of the three measures falls below cutoff. The number of patients identified on each measure is shown at the left; the three measures together identify 18 of 45 patients (some are below cutoff on more than one). (B) Prevalence of residual deficit under the primary recovery threshold (WAB-AQ ≥ 93.8, n = 45) and the stricter threshold used by Senthilkumar et al. (2026) (WAB-AQ ≥ 96.7, n = 33). Bars are point estimates; error bars are 95% confidence intervals.

### 3.2 Subgroup characteristics

Table 1 reports descriptive comparisons of subtle aphasia and no-deficit in WAB-recovered patients. Although there were no differences between groups in any one variable, the composite z-score separated the groups by construction (t(28.8) = −6.17, p < .001).

### 3.3 Differential capture across language measures

Figure 2 shows the overlap structure among the three additional language measures within the 18 subtle-deficit patients (Panel A) and the sensitivity of each measure subset for detecting these deficits (Panel B). The BNT alone captured 13 of 18 subtle-deficit patients (72%); the MCT CU count alone captured 11 of 18 (61%); the HANA alone captured 3 of 18 (17%). The two-measure protocol of the BNT plus the MCT content-unit count captured 18 of 18 subtle-deficit patients (100%), matching the all-three-measure protocol. The HANA contributed no patients that were not already flagged by either the BNT or the MCT CU count. Table 2 summarizes what each measure examines, why it adds information beyond the WAB-AQ, and its yield in the present sample.

**Figure 2.**
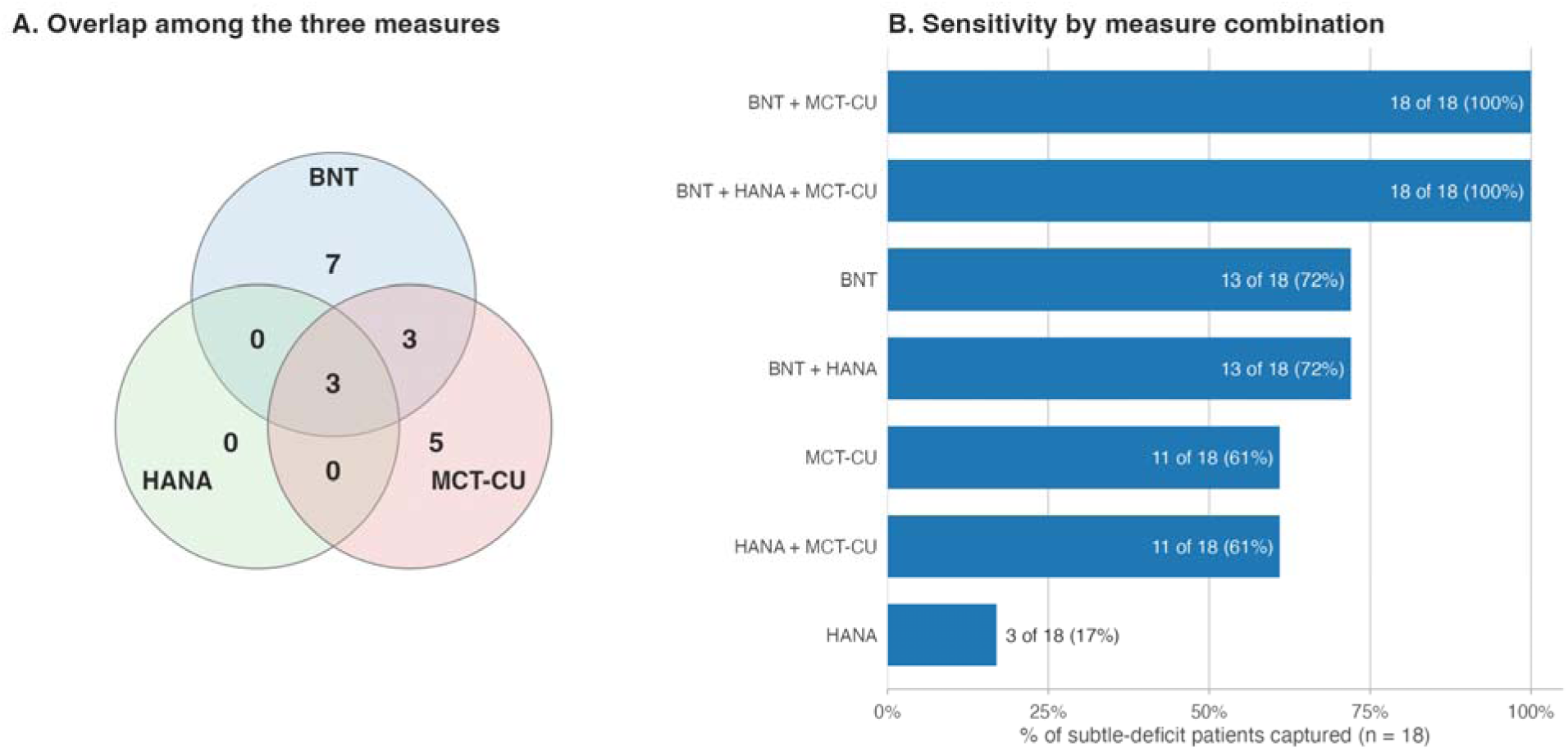
Differential capture of subtle language deficits within the WAB-recovered subgroup. Eighteen of 45 WAB-recovered patients (40%) were positive on at least one of the three language measures. (A) Venn diagram of the overlap structure among the three measures within this subtle-deficit set; region labels are patient counts. (B) Sensitivity of each non-empty subset of the three measures for detecting subtle deficits, sorted by capture rate. Bar labels are raw count and percentage of the 18 subtle-deficit patients.

**Table 2.** Language measures beyond the standard WAB-R Aphasia Quotient.

| Measure | Construct | Rationale | Admin (min) | Yield |
| --- | --- | --- | --- | --- |
| WAB-AQ ≥ 96.7 reading | Higher threshold on the standard WAB-R | Captures borderline-recovered patients whose AQ falls between 93.8 and 96.7 (Senthilkumar et al., 2026); the AQ is already obtained as part of standard care | 0 (re-read) | 12 of 45 (27%) reclassified out of recovered; captures 8 of 18 (44%) subtle-aphasia patients |
| Boston Naming Test (BNT-30 item form) | Single-word object naming with graded item difficulty | The WAB-R naming subtest is brief and easier; the BNT samples a wider object range with education-corrected normative cutoffs (Jefferson et al., 2007) | ~10 mins | 13 of 44 (30%) below education-corrected cutoff |
| Hopkins Action Naming Assessment (HANA) | Single-word verb / action naming | Verb naming dissociates from object naming; BNT and the WAB-R naming subtest do not capture action-specific deficits documented in stroke and PPA (Breining et al., 2022) | ~10 mins | 3 of 39 (8%) at or below the impairment cutoff |
| Modern Cookie Theft content units (MCT-CU) | Discourse-level propositional content in connected speech | Single-word tasks provide lexical scaffolding; connected speech reveals retrieval and content-generation deficits that escape word-level testing (Berube et al., 2019) | ~1-5 mins | 11 of 40 (28%) below the content-unit cutoff |

*Note.* Measures listed are those evaluated in or directly referenced by the present analyses. WAB-R = Western Aphasia Battery-Revised; AQ = Aphasia Quotient; Boston Naming Test = BNT; Hopkins Action Naming Assessment = HANA; Modern Cookie Theft = MCT; Content units = CU.

### 3.4 Correlate analyses within the recovered subgroup

#### 3.4.1 Hierarchical logistic regression on residual deficit (binary outcome)

Table 3 reports the full hierarchical logistic regression on the binary deficit indicator within the WAB-recovered subgroup. At Step 1 with aphasia history and sex, neither predictor was significant (aphasia history OR = 1.90, 95% CI [0.52, 6.92], p = .328; sex OR = 0.85 [0.23, 3.11], p = .809; AUC = 0.58, 95% CI [0.52, 0.76]). At Step 2, education’s contribution was marginal as an individual coefficient (OR = 0.81 per year [0.65, 1.01], p = .056) but the likelihood-ratio test against Step 1 was significant (LR χ²(1) = 3.98, p = .046; AUC = 0.69 [0.59, 0.88]). At Step 3, the addition of age did not improve fit (LR χ²(1) = 0.37, p = .543; AUC = 0.69 [0.62, 0.89]); age was non-significant (OR = 1.02 per year [0.96, 1.07], p = .546), and education became nominally significant as an individual coefficient (OR = 0.80 per year [0.64, 1.00], p = .049). At Step 4, the addition of composite residualized lesion load did not improve fit (LR χ²(1) = 0.12, p = .725; AUC = 0.69 [0.62, 0.91]) and the lesion coefficient was non-significant (OR = 0.92 [0.56, 1.50], p = .726). At Step 5, the addition of acute PHQ-9 produced a marginal improvement in fit (LR χ²(1) = 3.75, p = .053; AUC = 0.80, 95% CI [0.70, 1.00]); no individual coefficient reached significance, with PHQ-9 itself non-significant (OR = 1.25 per point [0.98, 1.60], p = .074) and education marginal (OR = 0.76 per year [0.58, 1.01], p = .057). The fully adjusted Step 5 binary model achieved discriminative accuracy at AUC = 0.80 (95% CI [0.70, 1.00]), with each step contributing incremental improvement from Step 1 (AUC = 0.58) through Step 5 (Figure 3, Panels A and C).

**Figure 3.**
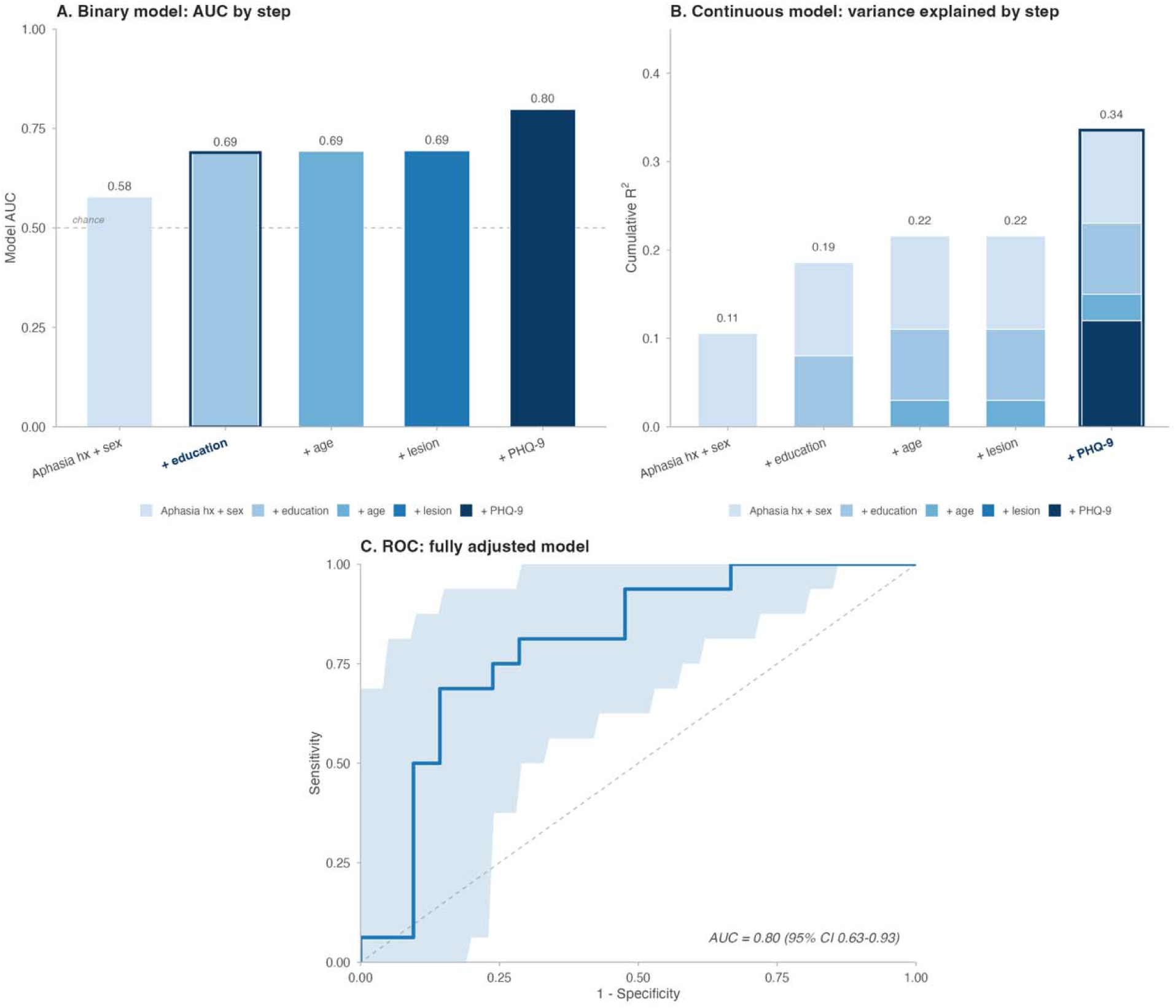
Hierarchical regression model summaries and ROC performance for the binary and continuous outcomes within the WAB-recovered subgroup. Panel A displays the per-step likelihood-ratio χ²(1) for each predictor block added to the binary logistic regression in the harmonized hierarchical order: aphasia history + sex (Step 1), + education (Step 2), + age (Step 3), + composite residualized lesion load (Step 4), + acute PHQ-9 (Step 5); bar heights show LR χ²(1) for each added block, and the overlaid line shows the running model AUC across steps. Panel B displays the cumulative variance explained in the continuous regression on the composite z-score across the same harmonized hierarchical order; stacked bar segments show each step’s incremental contribution, and the numeric label above each bar gives cumulative R² through that step. Panel C shows the receiver-operating-characteristic (ROC) curve for the full six-predictor binary logistic model within the WAB-recovered subgroup. Shaded bands are pointwise 95% bootstrap confidence intervals for sensitivity at fixed specificities. Diagonal dashed lines represent chance discrimination. AUC point estimates with bootstrap 95% confidence intervals are annotated on Panels A and C.

**Table 3.** Hierarchical logistic regression on residual deficit status (binary outcome) within the WAB-recovered subgroup.

| Step | n | Predictor | OR | 95% CI | P |
| --- | --- | --- | --- | --- | --- |
| Step 1 | 45 | Aphasia history | 1.90 | [0.52, 6.92] | .328 |
|  |  | Sex (female) | 0.85 | [0.23, 3.11] | .809 |
|  |  | AUC = 0.58 [0.52, 0.76] |  |  |  |
| Step 2 | 45 | Aphasia history | 1.75 | [0.45, 6.79] | .419 |
|  |  | Sex (female) | 0.71 | [0.18, 2.75] | .617 |
|  |  | Education (per year) | 0.81 | [0.65, 1.01] | .056 |
| | | LR $\chi^2(1) = 3.98$, $p = .046$;<br>AUC = 0.69 [0.59, 0.88] | | | |
| Step 3 | 45 | Aphasia history | 1.61 | [0.40, 6.43] | .503 |
|  |  | Sex (female) | 0.64 | [0.16, 2.63] | .540 |
|  |  | Education (per year) | 0.80 | [0.64, 1.00] | .049 |
|  |  | Age (per year) | 1.02 | [0.96, 1.07] | .546 |
| | | LR $\chi^2(1) = 0.37$, $p = .543$;<br>AUC = 0.69 [0.62, 0.89] | | | |
| Step 4 | 45 | Aphasia history | 1.51 | [0.29, 7.97] | .627 |
|  |  | Sex (female) | 0.51 | [0.12, 2.13] | .356 |
|  |  | Education (per year) | 0.81 | [0.64, 1.02] | .070 |
|  |  | Age (per year) | 1.01 | [0.96, 1.07] | .593 |
|  |  | Lesion-load composite | 0.92 | [0.56, 1.50] | .726 |
| | | LR $\chi^2(1) = 0.12$, $p = .725$;<br>AUC = 0.69 [0.62, 0.91] | | | |
| Step 5 | 37 | Aphasia history | 2.43 | [0.30, 19.77] | .405 |
|  |  | Sex (female) | 0.57 | [0.11, 3.09] | .517 |
|  |  | Education (per year) | 0.76 | [0.58, 1.01] | .057 |
|  |  | Age (per year) | 1.02 | [0.95, 1.10] | .599 |
|  |  | Lesion-load composite | 0.91 | [0.50, 1.64] | .747 |
|  |  | Acute PHQ-9 (per point) | 1.25 | [0.98, 1.60] | .074 |
| | | LR $\chi^2(1) = 3.75$, $p = .053$;<br>AUC = 0.80 [0.70, 1.00] | | | |

*Note.* OR = odds ratio with 95% confidence interval; LR $\chi^2$ = likelihood-ratio test against the previous nested step; AUC = area under the receiver operating characteristic curve with 95% bootstrap percentile confidence interval (2,000 resamples).

#### 3.4.2 Hierarchical linear regression on the continuous composite

Table 4 reports the parallel hierarchical linear regression on the continuous composite z-score within the WAB-recovered subgroup (n = 45 base; n = 37 at Step 5 owing to acute PHQ-9 missingness). At Step 1, acute aphasia history was associated with composite performance (β = −0.77, 95% CI [−1.40, −0.14], p = .018); the sex coefficient was small and non-significant (β = −0.13 [−0.75, 0.50], p = .688), and the model accounted for 13% of variance. At Step 2, the addition of education contributed significantly (ΔR² = 0.114; F-change p = .017) with education an independent positive correlate of the composite (β = 0.11 per year, 95% CI [0.02, 0.21], p = .017). At Step 3, the addition of age did not improve fit (ΔR² = 0.018; F-change p = .332); aphasia history remained significant (β = −0.64, p = .040), and education’s individual coefficient was significant (β = 0.12 per year, 95% CI [0.03, 0.21], p = .013). At Step 4, the addition of composite residualized lesion load produced essentially no change (ΔR² = 0.000; lesion coefficient = −0.01 [−0.23, 0.20], p = .894); aphasia history attenuated to non-significance once lesion load was included (β = −0.58, p = .124). At Step 5, the addition of acute PHQ-9 produced a substantial improvement (ΔR² = 0.178; F-change F(1, 30) = 8.60, p = .006; cumulative R² = 0.378), showing that acute depression independently predicted the composite (β = −0.13 per point, 95% CI [−0.22, −0.04], p = .006). However, we excluded a single outlier (acute PHQ-9 = 14, composite z = −2.80), and the outlier-excluded Step-5 PHQ-9 coefficient showed β = −0.056, 95% CI [−0.164, +0.051], p = .29, ΔR² = 0.027. In the Step 5 model, aphasia history (β = −0.69 [−1.49, 0.11], p = .087) and education (β = +0.08 per year [−0.02, 0.18], p = .101) were directionally consistent with earlier reduced models but did not reach significance (Figure 3, Panel B).

**Table 4.**
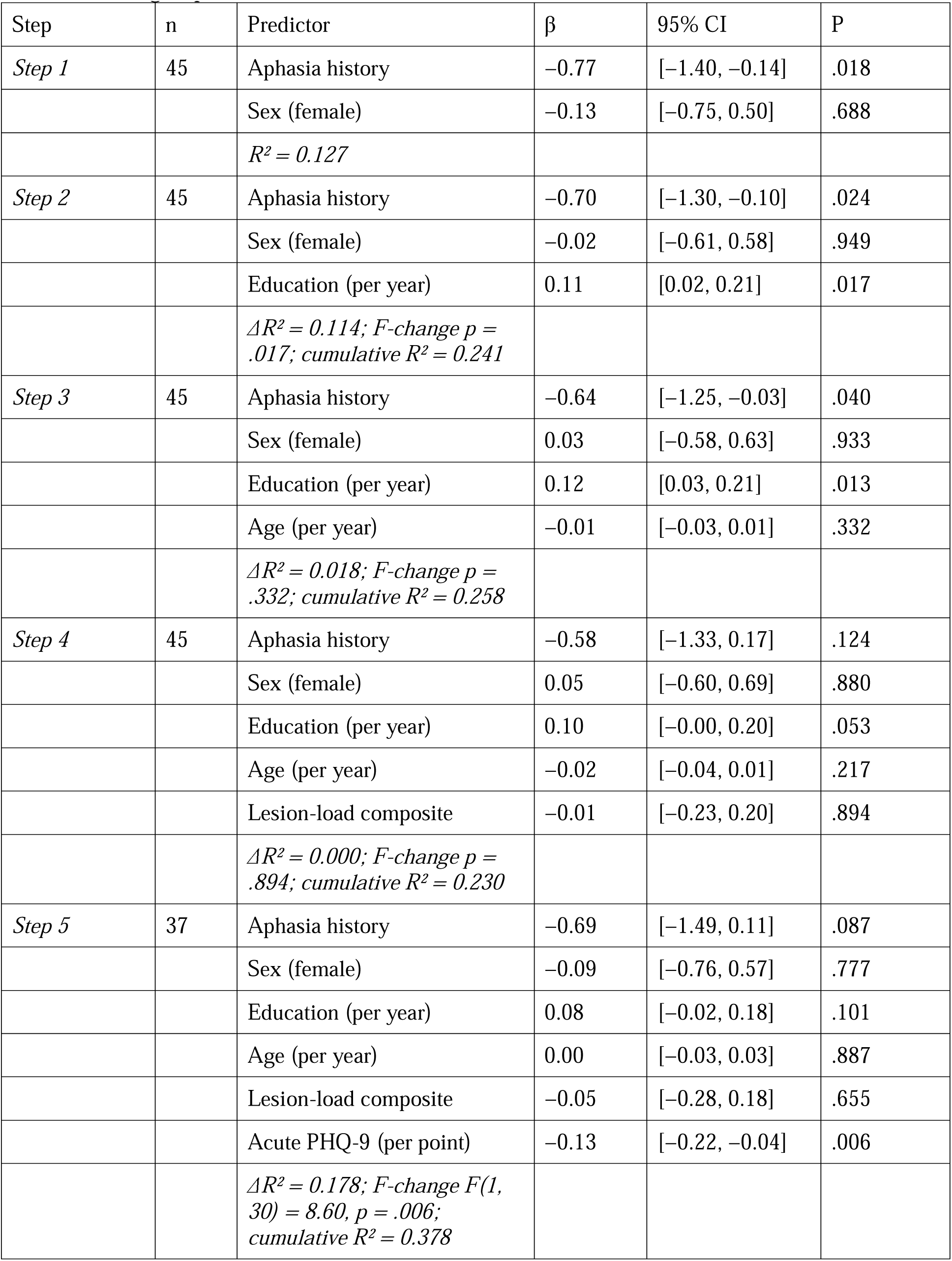
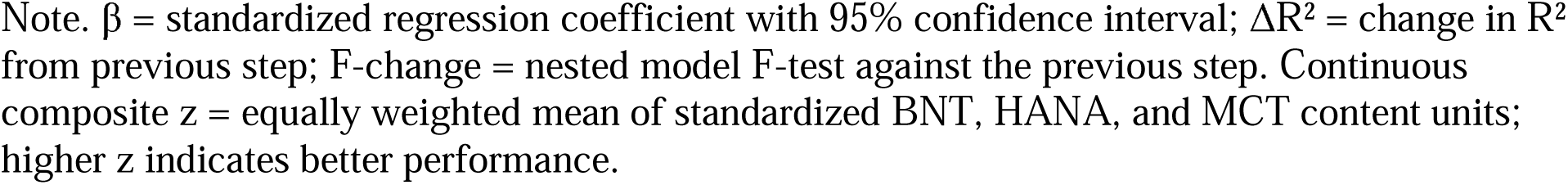
Hierarchical linear regression on continuous composite z-score within the WAB-recovered subgroup.

| Step | n | Predictor | $\beta$ | 95% CI | P |
| --- | --- | --- | --- | --- | --- |
| *Step 1* | 45 | Aphasia history | -0.77 | [-1.40, -0.14] | .018 |
|  |  | Sex (female) | -0.13 | [-0.75, 0.50] | .688 |
| | | $R^2 = 0.127$ | | | |
| *Step 2* | 45 | Aphasia history | -0.70 | [-1.30, -0.10] | .024 |
|  |  | Sex (female) | -0.02 | [-0.61, 0.58] | .949 |
|  |  | Education (per year) | 0.11 | [0.02, 0.21] | .017 |
| | | $\Delta R^2 = 0.114$; *F-change* $p = .017$; *cumulative* $R^2 = 0.241$ | | | |
| *Step 3* | 45 | Aphasia history | -0.64 | [-1.25, -0.03] | .040 |
|  |  | Sex (female) | 0.03 | [-0.58, 0.63] | .933 |
|  |  | Education (per year) | 0.12 | [0.03, 0.21] | .013 |
|  |  | Age (per year) | -0.01 | [-0.03, 0.01] | .332 |
| | | $\Delta R^2 = 0.018$; *F-change* $p = .332$; *cumulative* $R^2 = 0.258$ | | | |
| *Step 4* | 45 | Aphasia history | -0.58 | [-1.33, 0.17] | .124 |
|  |  | Sex (female) | 0.05 | [-0.60, 0.69] | .880 |
|  |  | Education (per year) | 0.10 | [-0.00, 0.20] | .053 |
|  |  | Age (per year) | -0.02 | [-0.04, 0.01] | .217 |
|  |  | Lesion-load composite | -0.01 | [-0.23, 0.20] | .894 |
| | | $\Delta R^2 = 0.000$; *F-change* $p = .894$; *cumulative* $R^2 = 0.230$ | | | |
| *Step 5* | 37 | Aphasia history | -0.69 | [-1.49, 0.11] | .087 |
|  |  | Sex (female) | -0.09 | [-0.76, 0.57] | .777 |
|  |  | Education (per year) | 0.08 | [-0.02, 0.18] | .101 |
|  |  | Age (per year) | 0.00 | [-0.03, 0.03] | .887 |
|  |  | Lesion-load composite | -0.05 | [-0.28, 0.18] | .655 |
|  |  | Acute PHQ-9 (per point) | -0.13 | [-0.22, -0.04] | .006 |
| | | $\Delta R^2 = 0.178$; *F-change* $F(1, 30) = 8.60$, $p = .006$; *cumulative* $R^2 = 0.378$ | | | |

Note. $\beta$ = standardized regression coefficient with 95% confidence interval; $\Delta R^2$ = change in $R^2$ from previous step; F-change = nested model F-test against the previous step. Continuous composite z = equally weighted mean of standardized BNT, HANA, and MCT content units; higher z indicates better performance.

#### 3.4.3 Convergent acute-chronic correlations

Acute scores on each of the three measures correlated with the 12-month continuous composite at moderate strength: acute BNT, r = .39 (n = 43, 95% CI [0.10, 0.62], p = .011); acute HANA, r = .38 (n = 40, 95% CI [0.07, 0.62], p = .016); acute MCT content units, r = .36 (n = 39, 95% CI [0.05, 0.61], p = .025). Speech-language therapy dose between acute and 12-month assessments did not correlate with the composite (r = −0.15, p = .375) or with binary deficit status (r = +0.18, p = .349). Bivariate Pearson correlations among all predictors and outcomes within the recovered subgroup are shown in Figure 4; the full correlation table is reported in Supplementary Table S2.

**Figure 4.**
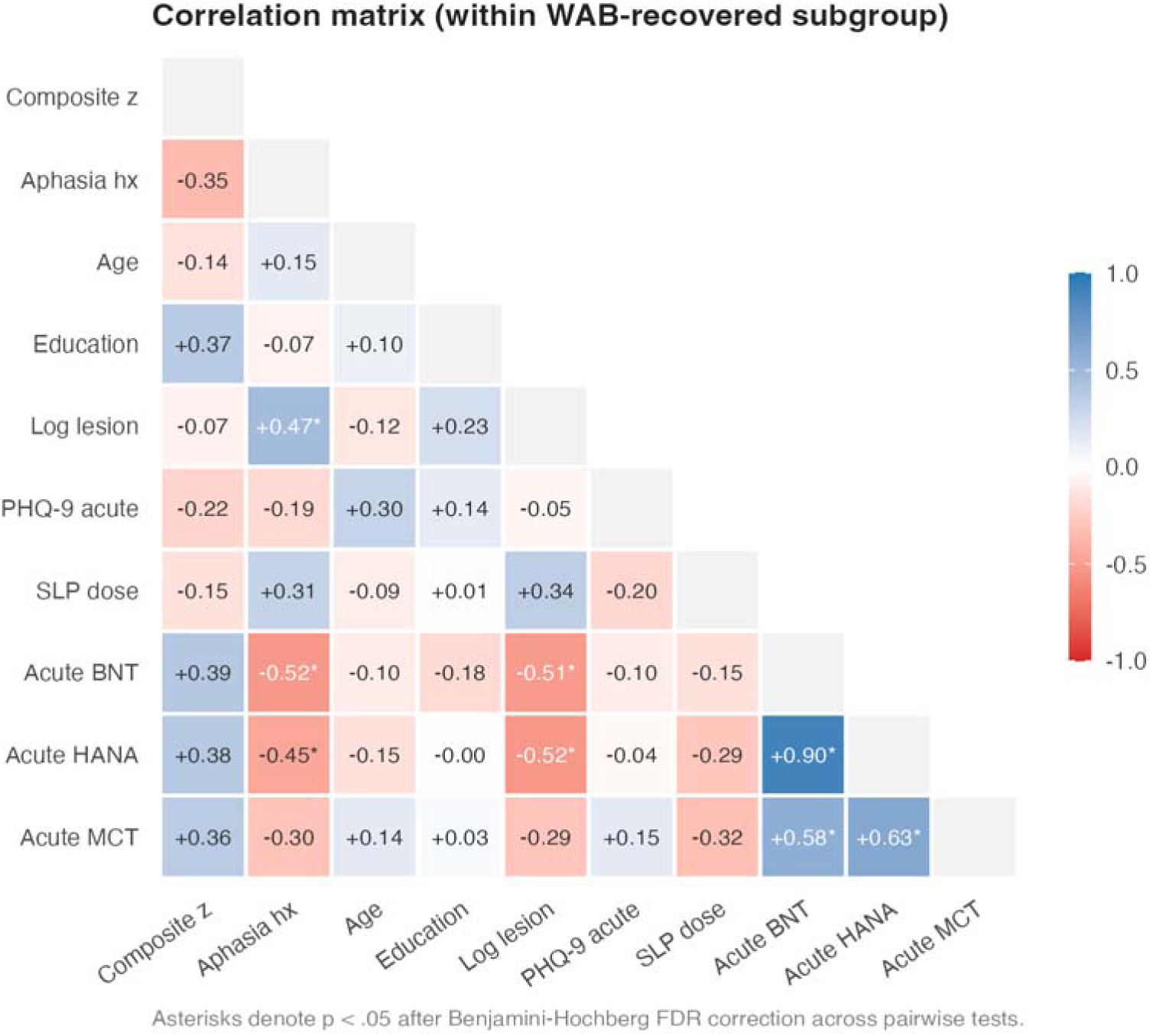
Pearson correlations among acute and 12-month variables within the WAB-recovered subgroup. Lower-triangle heatmap of pairwise Pearson correlations among the 12-month composite z-score, demographic and clinical covariates (aphasia history, age at stroke, years of education, log-transformed lesion volume, acute PHQ-9, total SLP sessions across the first 12 months), and acute language scores (BNT, HANA, MCT content units), computed across the WAB-recovered subgroup (n = 45; pairwise complete observations). Cell color encodes the Pearson coefficient on a diverging red-white-blue scale, with red indicating negative correlations and blue indicating positive correlations. Asterisks denote pairs with p < .05 after Benjamini-Hochberg false-discovery-rate correction across all 45 unique pairwise tests.

### 3.5 Structural lesion analyses

Composite residualized regional lesion load did not differ between deficit and no-deficit recovered patients (t(39.6) = −1.14, p = .260). Recall that when composite residualized lesion load was added at Step 4 of either the binary or the continuous regression model, it did not improve fit and the coefficient was non-significant (binary Step 4: OR = 0.92, p = .726; continuous Step 4: β = −0.01, p = .894).

## 4. Discussion

This study examined the prevalence and correlates of subtle language deficits at 12 months after left-hemisphere ischemic stroke. The findings adjudicate the three hypotheses introduced at the close of the Introduction. We hypothesized (H1) that residual deficits (subtle aphasia) would be detectable in a substantial minority of WAB-recovered patients. This hypothesis was supported: 18 of 45 WAB-recovered patients (40%) showed a residual deficit on at least one of the three additional language measures, approximating the range observed in prior literature. The differential measure analysis additionally showed that the Boston Naming Test and the Modern Cookie Theft content-unit count identified all 18 subtle deficit patients, whereas the Hopkins Action Naming Assessment was redundant with these two measures in the present sample. We hypothesized (H2) that demographic variables and lesion volume (but not regional lesion load) would differentiate WAB-recovered patients with subtle aphasia from those without. This hypothesis was partially supported. Acute self-reported depressive symptom severity (PHQ-9) emerged as a significant independent predictor in the fully adjusted continuous model, though sensitivity analyses indicated this continuous outcome association did not survive removal of a single high-influence observation; in the binary deficit model, the PHQ-9 block produced only a marginal improvement in fit and the individual coefficient did not reach significance; educational attainment was an independent correlate of the continuous composite and a borderline-significant predictor of binary deficit status; and acute aphasia history was a significant correlate in reduced models of the continuous outcome but attenuated to non-significance once composite residualized lesion load was included. Furthermore, contrary to the hypothesized role of lesion volume, neither gross lesion volume nor composite residualized regional lesion load differentiated the groups, whether at the subgroup level or as a covariate in either regression model. Lastly, we hypothesized (H3) that application of the Senthilkumar et al. (2026) alternative recovery cutoff (WAB-AQ ≥ 96.7) would capture some WAB-recovered patients with subtle aphasia, indicating that an alternative threshold can address the upper-end sensitivity problem. This hypothesis was largely supported. Application of the stricter ≥96.7 cutoff reclassified 12 of 45 WAB-recovered patients (27%) out of recovered status; within the remaining still-recovered subgroup (n = 33), 10 patients still showed a residual deficit. The stricter threshold addresses part of the upper-end sensitivity problem by reclassifying borderline-recovered patients but does not eliminate the need for additional language testing to identify residual deficits that persist within the still-recovered subgroup.

The 40% prevalence we identified adds to a literature in which prevalence estimates rely on heterogeneous denominators and detection methods. Conservative estimates cluster at 11-19%: Kurland et al. (2026), in n = 96 BATS participants with self-identified aphasia at intake, identified 17% as not-aphasic-by-WAB whose continued self-perceived communicative difficulty was quantified post-hoc using the Aphasia Communication Outcome Measure (ACOM; Hula et al., 2015); Senthilkumar et al. (2026), in n = 171 acute LH stroke patients, identified 19% with discourse-level aphasia despite a non-aphasic WAB-R score. Discourse-corpus estimates of NABW status fall on the lower end of this range (Fromm et al., 2024, 11% NABW in n = 281 AphasiaBank PWA). Two features of the present design likely contribute to the higher estimate. First, multi-measure detection (BNT, HANA, MCT) is more sensitive than any single-measure approach; the differential assessment analysis confirmed that the Boston Naming Test and Modern Cookie Theft content-unit count jointly identified all subtle-deficit cases in the present sample, whereas approaches relying on a single discourse or functional-communication measure would necessarily miss naming-specific deficits that confrontation testing elicits. Second, the deficit-classification rule is sensitive by construction, treating any below-threshold measure as evidence of residual deficit. Importantly, the gap is unlikely to be primarily attributable to denominator differences: when the stricter Senthilkumar et al. (2026) cutoff (≥96.7) was applied within the present sample (Figure 1, Panel B), residual-deficit prevalence in the still-recovered subgroup remained well above the 11-19% range, suggesting that the higher estimate reflects detection sensitivity rather than a more permissive recovery threshold. One future direction should involve developing collaborative, consensus-driven approaches to the principled identification of residual or subtle deficits, including which sensitive measures should constitute a minimum detection protocol.

Next, in the fully adjusted continuous model, higher acute PHQ-9 predicted poorer 12-month composite language performance, and the PHQ-9 block contributed the largest variance increment among the five predictor blocks; however, the association proved sensitive to a single high-influence observation. With that patient removed, the PHQ-9 association was no longer significant and the Step-5 variance increment shrank substantially, indicating that the depression-language association should be interpreted with caution at the present sample size. Even so, three non-mutually-exclusive accounts remain. First, subclinical residual language impairment may generate communicative frustration that drives mood symptoms, in that the language deficit precedes and partly produces the depressive presentation, consistent with longitudinal evidence that communication impairment and aphasia severity predict subsequent emotional distress and depressive symptoms (Hilari et al., 2010, 2012). Second, acute depressive symptoms may impair the cognitive and motivational resources that support performance on sensitive language measures, consistent with evidence that post-stroke depression is associated with reduced functional and cognitive recovery (Kutlubaev & Hackett, 2014; Robinson & Jorge, 2016). Third, mood symptoms and language performance may share a substrate of frontal and limbic dysfunction without one directly causing the other, consistent with engagement of domain-general cognitive-control networks in language recovery from aphasia (Brownsett et al., 2014; Geranmayeh et al., 2014). Recent evidence tenuously supports this third reading: Marte et al., 2026, in 57 chronic PWA and 43 healthy controls, found that a composite measure of language and emotion deficits in chronic aphasia (derived from naturalistic movie-viewing tasks) was independently associated with both WAB-AQ and PHQ-8 scores, even though WAB-AQ and PHQ-8 themselves were not significantly correlated, a dissociation the authors interpreted as evidence that depression in aphasia is driven by factors beyond language deficit per se, including independent emotion-processing impairments that may share a frontal-network substrate with language production. The present finding that acute PHQ-9 predicted the 12-month composite even when acute clinical, demographic, and structural variables were modeled together is consistent with the second and third accounts; however, the present data are cross-sectional with respect to mood and outcome (i.e., we did not measure change in PHQ-9 scores over time), the PHQ-9 effect did not survive outlier removal, hence the available evidence cannot adjudicate between these accounts. A further nuance the present aggregate analysis cannot resolve is that the three language measures need not relate to mood uniformly, and that the PHQ-9 combines affective items with items indexing concentration and psychomotor slowing that can be endorsed for reasons partly entangled with language and cognitive performance itself. Whether the mood-language association is uniform across confrontation naming, action naming, and connected-speech content, and how it partitions across affective versus cognitive symptom items, are questions that measure- and item-level analyses are better positioned to address.

Acute aphasia history was a significant correlate in Steps 1-3 of the continuous regression but attenuated to non-significance once composite residualized lesion load was added at Step 4, and remained so through the fully adjusted Step 5 model. This pattern is consistent with the aphasia history variance being largely taken by the lesion load composite, since acute aphasia history is itself an indirect index of stroke severity and lesion burden. The substantive finding from the earlier steps, that more severe acute language impairment predicts poorer 12-month outcome, is therefore not negated, rather, its independent contribution above and beyond demographic, structural, and mood covariates does not reach significance at the present sample size. This finding is reflected in recent recovery literature in which demographic and reserve-linked variables account for significant outcome variance alongside acute severity once these are entered into models (Johnson et al., 2022).

Further, education emerged as a correlate of subtle aphasia, an independent positive predictor of the continuous composite and a borderline-significant predictor of binary deficit status at Step 3 of both regressions (Tables 3 and 4). The education contribution admits at least two competing interpretations. First, on a cognitive reserve account (Stern, 2009; 2012; Umarova et al., 2021), education indexes pre-stroke neural and cognitive resources that buffer the functional expression of brain injury. Once the structural anatomical threshold for recovery has been met, what separates patients with and without residual deficits may be the compensatory capacity brought to the injury rather than the amount of tissue destroyed. On this view, what education indexes is the pre-stroke neural architecture available to support compensation, a latent property statistically detectable in behavioral proxies like education and not (or not fully) detectable in focal-lesion metrics. Consistent with this account, several studies have reported that measures of preserved network architecture accounted for variance in chronic outcomes above and beyond focal damage (Erickson et al., 2022; Gleichgerrcht et al., 2015; Griffis et al., 2020). In other words, the brain’s residual network organization, which education is thought to shape (Bathelt et al., 2019), contributes to recovery beyond what lesion location and volume can explain. Second, education may proxy for pre-stroke lexical-semantic and discourse-level abilities, such that below-cutoff performance on the present language battery partly reflects pre-stroke rather than post-stroke variation in those abilities. This concern is partly addressed by the education-corrected BNT cutoff (Jefferson et al., 2007), but the HANA and MCT content-unit cutoffs are not education-corrected; while these cutoffs were derived from comparable control populations, they cannot fully equate pre-stroke language ability across individual patients. The two accounts are not mutually exclusive as well, pre-stroke language ability and pre-stroke cognitive reserve are themselves correlated (Stern, 2009, 2012; Ali et al., 2021; González-Fernández et al., 2011; Umarova et al., 2021), and disentangling them within the present cross-sectional, post-stroke-only design is not possible.

Next, above the WAB-R recovery threshold, residualized regional lesion load carries little independent discriminative information, and the composite did not differentiate deficit and no-deficit recovered patients in any adjusted analysis, and adding it to either regression failed to improve discriminative accuracy. The likely mechanism is selection on outcome, in that patients with large lesions are disproportionately in the persistent aphasia group (Bonkhoff et al., 2022), leaving reduced anatomical variance within the recovered subgroup and limiting focal measures’ statistical power at the upper performance range. Convergent evidence supports this interpretation, e.g., (Bunker et al., 2025), in a partly overlapping cohort, reported small lesion volumes in the WAB-recovered (NABW) subgroup relative to the aphasic subgroup. The recovered range may therefore be a subgroup in which voxel-based focal-damage metrics are underpowered and, potentially, connectivity-, microstructure-, or brain health-based methods are better suited.

Clinically, the present findings support the position that the WAB-AQ alone is insufficient evidence of complete language recovery, particularly when patients carry markers of elevated risk. A clinician evaluating a patient who scores at or above the recovery cutoff should consider that fewer years of formal education and an acute aphasia history increase risk of residual language deficits detectable on multi-measure assessment, and elevated acute depressive symptoms may compound this risk and is independently important to address (Harmon, 2024). Brief sensitive measures such as the SCANT (Walker et al., 2022) or rapid letter fluency (Laks et al., 2025; 2026) would identify a sizable fraction of recovered patients as well.

This study has several limitations. Conceptually, “subtle aphasia” as operationalized here is a measure-and-cutoff-dependent construct rather than a discrete clinical category, and prevalence figures reflect detection on the present battery with the present cutoffs, and an alternative sensitive battery could yield substantially different estimates. The 12-month assessment is also a single snapshot, and whether the identified deficits remit, persist, or worsen over subsequent years is beyond the present design, though an interesting and important future direction. The link between test-based residual deficit and real-world communicative function is similarly not directly examined, and whether the subtle deficit subgroup reports or exhibits everyday communicative difficulty is an empirical question the present data do not address.

Generalizability is further bounded by the acute LH ischemic stroke focus and English-speaking enrollment, and the construct may differ in e.g., right-hemisphere, hemorrhagic, bilateral, or non-English-speaking populations. Technically, connectivity-based, microstructural, or brain health approaches may detect what ROI-based lesion load did not. Lastly, the depression-language association is cross-sectional with respect to both variables, so longitudinal mood assessment paired with mediation or treatment-controlled designs would be needed to identify directional relationships.

## 5. Conclusions

The present study makes three contributions. Methodologically, it operationalizes subtle aphasia at the individual-patient level using published normative cutoffs, establishes a 40% subtle aphasia prevalence within the WAB-recovered subgroup, and identifies a two-measure protocol (the Boston Naming Test plus the Modern Cookie Theft content-unit count) as sufficient to capture all subtle deficit patients in the present sample. Empirically, it identifies educational attainment as a significant correlate of residual deficit status, acute aphasia history as a significant correlate in certain models, and acute self-reported depressive symptoms (PHQ-9) as a candidate correlate that warrants confirmation in larger samples. Neuroanatomically, composite residualized regional lesion load did not differentiate subtle aphasia and no deficit recovered patients in any adjusted analysis.

Clinically, the present findings support the position that the WAB-AQ alone is insufficient evidence of complete language recovery, and that brief sensitive measures should be administered to recovered patients with an acute aphasia history, particularly if they have lower educational attainment, and that the role of elevated acute depressive symptoms is an important future direction in examining the full breadth of recovery.

## Ethical approval

All procedures were approved by the Johns Hopkins Institutional Review Board. All participants provided written informed consent in accordance with the Declaration of Helsinki.

## Data availability statement

Our data are available from the Vivli controlled repository using the Sponsor Protocol ID: NA_00042097 (https://doi.org/10.25934/PR00012002). The identifiable data may not be shared publicly, as they represent clinical protected health information in participants who did not consent to have their data banked. Currently, data are available upon request to the authors, subject to review by the Johns Hopkins University School of Medicine Institutional Review Board (contact via) resulting in a formal data sharing agreement.

## Author contributions

Manuel Jose Marte: Conceptualization, Methodology, Formal analysis, Investigation, Data curation, Visualization, Writing – original draft, Writing – review & editing. Mathew Chaves: Data curation, Writing – review & editing. Lindsey Kelly: Investigation, Writing – review & editing. Isidora Diaz-Carr: Investigation, Writing – review & editing. Voss Neal: Investigation, Writing – review & editing. Andreia V. Faria: Methodology, Software, Resources, Funding acquisition, Writing – review & editing. Melissa D. Stockbridge: Conceptualization, Funding acquisition, Supervision, Writing – review & editing. Argye E. Hillis: Conceptualization, Resources, Supervision, Project administration, Funding acquisition, Writing – review & editing.

## Acknowledgements

The authors sincerely thank the study participants and their families for their generous contribution of time, as well as research coordinators, speech-language pathologists who contributed to acute assessments, and our imaging technologists.

## Declaration of generative AI use

Generative AI (ChatGPT-5.5) was used to assist with editing manuscript text. The study design, statistical analyses, interpretation of results, and all conclusions are the authors’ own; no research data, results, figures, or references were generated by AI. The authors take full responsibility for the content and integrity of the work.

