## Supplemental Materials for "Subtle language deficits in WAB-recovered patients at 12 months after left-hemisphere stroke"

**Supplementary Table S1.** *Sample characteristics by 12-month WAB-AQ recovery group (N = 67).*

| Variable | Never aphasic (n = 30) | Resolved (n = 15) | Persistent (n = 22) | Test, p |
| --- | --- | --- | --- | --- |
| Age, years (M, SD) | 58.5 (11.2) | 62.5 (15.5) | 62.5 (11.4) | F = 0.78, p = .461 |
| Female sex, n (%) | 13 (43) | 3 (20) | 12 (55) | χ²(2) = 4.43, p = .109 |
| Education, years (M, SD) | 15.0 (3.1) | 14.5 (3.0) | 13.3 (2.3) | F = 2.13, p = .128 |
| Days post-onset (M, SD) | 2.4 (1.6) | 3.9 (2.4) | 3.3 (2.8) | F = 2.22, p = .117 |
| Acute WAB-AQ (M, SD) | 97.8 (1.7) | 72.8 (24.8) | 34.0 (31.5) | F = 92.4, p < .001 |
| 12-month WAB-AQ (M, SD) | 98.6 (1.2) | 96.1 (1.9) | 62.9 (32.2) | F = 31.5, p < .001 |
| Log lesion volume (M, SD) | 7.6 (1.4) | 9.2 (1.8) | 11.0 (1.1) | F = 41.3, p < .001 |

*Note. Mean (SD) shown for continuous variables; n (%) for categorical. Continuous variables compared with Welch one-way ANOVA across the three groups; female sex compared with Pearson chi-square. The recovered analytic subgroup (n = 45) combines the never-aphasic (n = 30) and resolved-aphasia (n = 15) groups. WAB-AQ = Western Aphasia Battery–Revised Aphasia Quotient.*

**Supplementary Table S2.** *Bivariate Pearson correlations within the WAB-recovered subgroup.*

| Predictor | Outcome | n | Pearson r | 95% CI (p) |
| --- | --- | --- | --- | --- |
| Acute BNT-30 | Composite z (12 months) | 43 | +.39 | [+.10, +.62] (p = .011) |
| Acute HANA | Composite z (12 months) | 40 | +.38 | [+.07, +.62] (p = .016) |
| Acute MCT content units | Composite z (12 months) | 39 | +.36 | [+.05, +.61] (p = .025) |
| SLP sessions, 0–12 months | Composite z (12 months) | 37 | −.15 | [−.45, +.18] (p = .375) |
| Acute PHQ-9 | Composite z (12 months) | 37 | −.39 | [−.63, −.07] (p = .018) |

*Note. Pearson r within the WAB-recovered subgroup. Composite z-score is the equally weighted mean of standardized BNT, HANA, and MCT content units. BNT = Boston Naming Test; HANA = Hopkins Action Naming Assessment; MCT = Modern Cookie Theft; SLP = speech-language therapy; PHQ-9 = Patient Health Questionnaire-9.*
